# Nationwide upsurge in invasive disease in the context of longitudinal surveillance of carriage and invasive *Streptococcus pyogenes* 2009-2023, the Netherlands: a molecular epidemiological study

**DOI:** 10.1101/2024.05.16.24307270

**Authors:** Lidewij W. Rümke, Matthew A. Davies, Stefan M.T. Vestjens, Boas C.L. van der Putten, Wendy C.M. Bril-Keijzers, Marlies A. van Houten, Nynke Y. Rots, Alienke J. Wijmenga-Monsuur, Arie van der Ende, Brechje de Gier, Bart J.M. Vlaminckx, Nina M. van Sorge

## Abstract

**Background:** Since 2022, many countries reported an upsurge in invasive group A streptococcal (iGAS) infections. We explored whether changes in *S. pyogenes* carriage rates or emergence of more virulent strains, such as *emm*1 variants M1_UK_ and M1_DK_, contributed to the 2022/2023 surge in the Netherlands.

**Methods:** We determined *emm* (sub)type distribution for 2,698 invasive and 351 *S. pyogenes* carriage isolates collected between January 2009 - March 2023. Genetic evolution of *emm*1 was analyzed by whole-genome sequencing of 497 *emm*1 isolates.

**Findings:** The nationwide iGAS upsurge coincided with a sharp increase of *emm*1.0 from 18% (18/100) of invasive isolates in Q1 2022 to 58% (388/670) in Q1 2023 (Fisher’s exact, p<0.0001). M1_UK_ became dominant among invasive *emm*1 isolates in 2016 and further expanded from 72% in Q1 2022 to 96% in Q1 2023. Phylogenetic comparison revealed evolution and clonal expansion of four new M1_UK_ clades in 2022/2023. DNase Spd1 and superantigen SpeC were acquired in 9% (46/497) of *emm*1 isolates. *S. pyogenes* carriage rates and *emm*1 proportions in carriage isolates remained stable during this surge and the expansion of M1_UK_ in iGAS was not reflected in carriage isolates.

**Interpretation:** During the 2022/2023 iGAS surge in the Netherlands, expansion of four new M1_UK_ clades was observed among invasive isolates but not carriage isolates, suggesting increased virulence and fitness of M1_UK_ compared to contemporary M1 strains. The emergence of more virulent clades has important implications for public health strategies such as antibiotic prophylaxis for close contacts of iGAS patients.

## Introduction

*Streptococcus pyogenes* (commonly referred to as Group A *Streptococcus*, GAS) accounts for an estimated 500,000 annual deaths worldwide, classifying this bacterial pathogen as one of the top ten causes of infectious disease-related mortality (1). The clinical spectrum ranges from asymptomatic carriage, non-invasive infections such as impetigo, pharyngitis and scarlet fever, to life-threatening invasive disease including (puerperal) sepsis, necrotizing fasciitis and progression to streptococcal toxic shock syndrome (STSS) (2). Several European countries and the United States reported a striking increase in invasive GAS (iGAS) infections after lifting of COVID-19-related social restrictions in 2022 (3, 4), with shifts in the relative contribution of specific invasive *S. pyogenes* disease manifestations compared to previous epidemiological observations (5–9).

Two years of COVID-19-related restriction measures have limited the transmission of and exposure to *S. pyogenes* and iGAS-predisposing viruses (e.g. varicella zoster virus, influenza virus) (10). It has been speculated that this reduced intermittent exposure may have caused a waning of population immunity (‘immunity debt’), resulting in an increased proportion of susceptible individuals. A previous study showed that varicella and respiratory virus infections only partly explained the iGAS increase (10). Also, iGAS incidences remained persistently higher than pre-COVID levels after viral infections declined, suggesting changes in bacterial characteristics. It is currently unclear whether new *S. pyogenes* variants or changes in recently-emerged *S. pyogenes* lineages, such as the toxicogenic M1_UK_ (11) and M1_DK_ variants (12), have contributed to the recent outbreak in the Netherlands.

For epidemiological purposes, *S. pyogenes* isolates are classified based on sequence variation of the *emm* gene (13), which encodes the surface-expressed virulence factor M protein. *Emm*1.0 has been a dominant *emm* type among strains causing a resurgence in iGAS infections in industrialized countries since the 1980s (14) due to the evolution of a highly epidemic clone referred to as M1T1 or M1_global_. M1_global_ acquired increased virulence due to three subsequent genetic events: the acquisition of two phages encoding for virulence factors DNase SdaD2 (Sda1) and scarlet fever toxin A (SpeA), and the uptake of a large chromosomal region encoding the toxins NAD glycohydrolase and streptolysin O (15, 16). In 2019, the emergence of a M1_global_ variant, named M1_UK_, was identified during a period of increased scarlet fever and iGAS activity in the United Kingdom (11). This new clone can be differentiated from M1_global_ by 27 lineage-defining single-nucleotide polymorphisms (SNP) and is characterized by increased SpeA production (11, 17), resulting from a SNP in the transfer-messenger RNA gene *ssrA* (15). Numerous countries, including the United States, Canada, the Netherlands, Australia, and Taiwan, have reported on the presence and expansion of the M1_UK_ lineage among iGAS patients, confirming global dissemination and further evolution of this new variant (15, 18–21).

In the Netherlands, three invasive clinical presentations have been notifiable by law since 2008: necrotizing fasciitis, STSS, and puerperal fever- or sepsis. Peak incidences of necrotizing fasciitis were seen in the first half of 2022 and a sharp increase of STSS in December 2022 (22). In addition, bacteriological surveillance covering all iGAS manifestations similarly detected exceptionally high numbers of iGAS cases, equating to an iGAS incidence of approximately 10 per 100,000 individuals/year. We hypothesized that the emergence of more virulent lineages could be underlying the nationwide sustained surge of iGAS disease. To this end, we analyzed shifts in *emm* type prevalence based on 2,698 invasive *S. pyogenes* strains and 351 isolates of asymptomatic carriers from the open population over an extended time period including years before, during and after the COVID-19 pandemic (2009–2023). In addition, 497 *emm*1 isolates were analyzed by whole-genome sequencing (WGS) to determine genetic diversification of *emm*1 in relation to the nationwide iGAS surge.

## Methods

### S. pyogenes strain collections

*S. pyogenes* strains were available from three collections (Table 1): A) national invasive isolates (January 2019 through March 2023) cultured from normally sterile sites or non-sterile sites with clinical iGAS (e.g. puerperal sepsis in combination with a vaginal swab), submitted since January 2019 to the Netherlands Reference Laboratory for Bacterial Meningitis (NRLBM, Amsterdam UMC, Amsterdam, The Netherlands) by nine medical microbiology laboratories covering ∼28% of the Dutch population. Since April 2022, all medical microbiology laboratories submitted isolates to the NRLBM; B) retrospectively collected and recultured invasive isolates obtained from blood- and cerebrospinal fluid (CSF) from patients admitted between January 2009 and December 2019 to the University Medical Center Utrecht (Utrecht), St. Antonius hospital (Utrecht, Nieuwegein) or Diakonessenhuis (Utrecht, Zeist); C) carriage isolates from naso- or oropharyngeal swabs collected as part of six pneumococcal carriage surveillance studies (OKIDOKI studies), conducted triennially between 2009 and 2023 in the Noord-Holland region (23).

**Table 1.** *S. pyogenes* strain collections from patients with invasive group A streptococcal disease and asymptomatic carriers in the Netherlands, 2009-2023.

| Strain collection | Region | Time period | Specimen of isolation | Age groups |
| --- | --- | --- | --- | --- |
| **A** Invasive national (n=2,426)<sup>a</sup> | Nationwide, Netherlands | Jan 2019 - Mar 2023 | Any sterile body compartment or non-sterile site with clinical iGAS | All ages |
| **B** Invasive regional (n=272)<sup>b</sup> | Utrecht region, Netherlands | Jan 2009 - Dec 2019 | Blood, CSF | All ages |
| **C** Carriage regional (n=351)<sup>c</sup> | North Holland region, Netherlands | Mar 2009 - Jul 2009<sup>d,e,g</sup><br>Sep 2010 - Jan 2011<sup>d,e,g</sup><br>Oct 2012 - Mar 2013<sup>d,e,g,h</sup><br>Oct 2015 - Feb 2016<sup>e,f,g</sup><br>Oct 2018 - Mar 2019<sup>e,g</sup><br>Oct 2022 - Feb 2023<sup>e,g</sup> | Naso- and oropharyngeal swabs | 11-, 24- and 46-month old, adults |

CSF, cerebral spinal fluid. <sup>a</sup>Invasive isolates, defined as cultured from a normally sterile site or a non-sterile site with clinical iGAS (e.g. puerperal sepsis in combination with a vaginal swab), submitted to the Netherlands Reference Laboratory for Bacterial Meningitis (NRLBM) by nine medical microbiological laboratories covering ~28% of the Dutch population. Since April 2022, all medical microbiology laboratories submitted invasive isolates; <sup>b</sup>Recovered invasive isolates (blood, CSF) from patients admitted to the University Medical Center Utrecht (Utrecht), St. Antonius hospital (Nieuwegein, Utrecht) or Diaconessenhuis (Utrecht, Zeist); <sup>c</sup>Carriage isolates collected as part of six pneumococcal carriage surveillance studies (OKIDOKI studies) conducted triennially; <sup>d</sup>11-month old children; <sup>e</sup>24-month old children; <sup>f</sup>46-month old children; <sup>g</sup>Parents of the 24-month old children; <sup>h</sup>Adults with limited contact with children aged ≤5 years

### Emm typing and whole genome sequencing of emm1 S. pyogenes strains

All strains were *emm*-genotyped by conventional PCR amplification and subsequent Sanger sequencing of the 180-bp hypervariable domain of the *emm* gene according to the CDC protocol. A subset of *emm*1 isolates was submitted for next-generation sequencing using Illumina short-read technology (Microbes NG, Birmingham, UK and in-house core facility Amsterdam UMC). Genomic analyses included extensive quality control with Trimmomatic, *de novo* assembly with SPAdes, resistance gene identification with Abricate, reference based read mapping and SNP calling with Snippy, and identification of recombinant regions with Gubbins and phylogenetic analysis with IQTree. Details of the methods, including relevant WGS parameters, software and an overview of the strains, are provided in the Supplementary material.

### Statistical analysis

Categorical variables were described as frequencies and percentages and compared using the Fisher’s exact test. Carriage rates are presented as point estimates with a binomial confidence interval. Binomial logistic regression was used to assess the association of *emm* type and invasiveness versus carriage with adjustment for time period and age group. Adjusted odds ratio’s (OR) and their 95% confidence intervals were calculated. *P*-values less than 0.05 were considered statistically significant. Statistical analyses were performed using SPSS software (version 27.0 for Windows; Chicago, Illinois, USA). Figures were made using GraphPad Prism (version 9.3.0).

### Ethics statement

Invasive *S. pyogenes* isolates were collected as part of routine care. Carriage isolates were collected after written informed consent. Four carriage surveillance studies were approved by acknowledged Dutch medical ethics committees (NL24116.000.08, NL40288.094.12; NL53027.094.15 NL65919.100.18), two carriage surveillance studies did not need ethical approval (one was registered in the trial registry under ISRCTN31549735). All were conducted in accordance with the European Statements for Good Clinical Practice and the Declaration of Helsinki of the World Medical Association. Only fully anonymized data was used.

## Results

Across the study period, national notifications of non-puerperal iGAS (STSS or necrotizing fasciitis) were markedly increased in 2022 (n=361) compared to pre-COVID-19 (annual mean 2009-2019: n=165) and two pandemic years (n=84 in 2020, n=51 in 2021). This steep increase was not reflected in *S. pyogenes* (naso)pharyngeal carriage rates, which fluctuated between 2%-13% (median 5%) in 24-month old children and 3%-5% (median 4%) in their parents between 2009-2019, compared to 7% in both groups in winter 2022-2023 (Figure 1).

**Figure 1.**
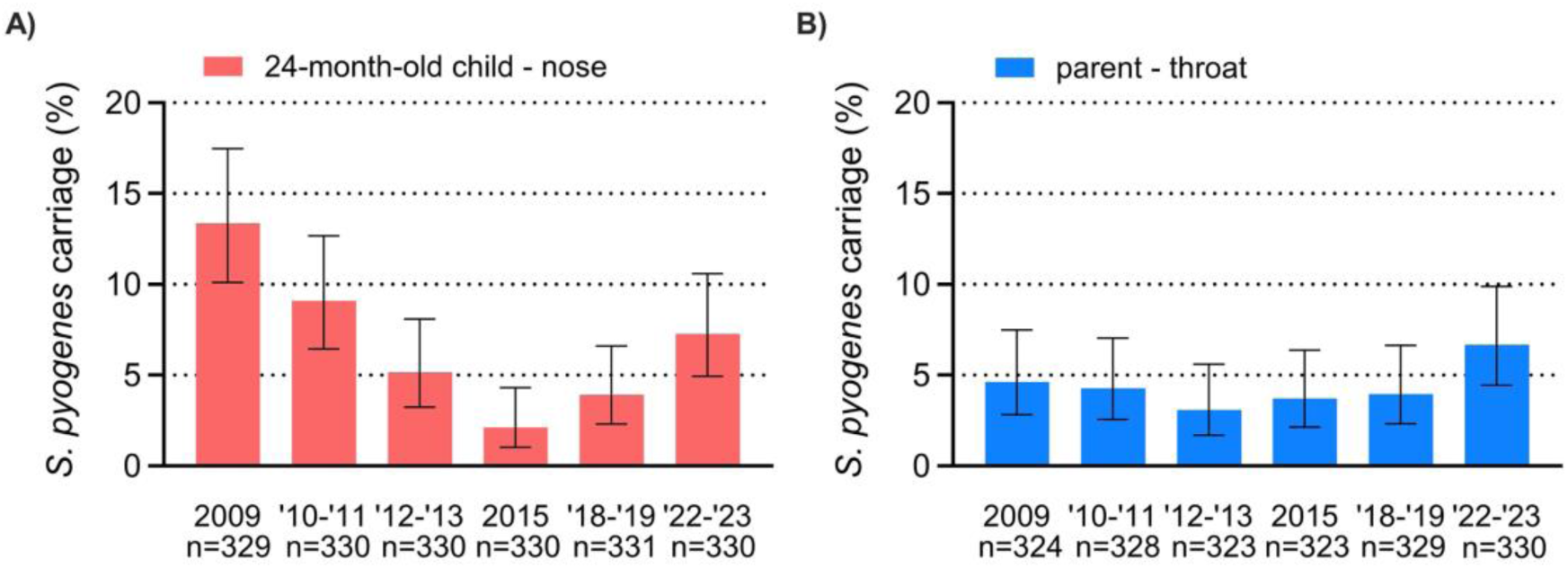
Asymptomatic nasopharyngeal *S. pyogenes* carriage rates in 24-month old children and their parents, 2009-2023. Percentage of *S. pyogenes* carriage in nasopharyngeal swabs of 24-month old children (A swabs) and pharyngeal swabs of their parents (B) collected in the Netherlands in the following time intervals: March 2009 through July 2009, September 2010 through January 2011, October 2012 through March 2013, October 2015 through February 2016, October 2018 through March 2019, and October 2022 through February 2023. Carriage rates are presented as point estimates with a binomial confidence interval. The numbers below the graph indicate denominators.

We next determined *emm* genotypes of 2,426 national invasive *S. pyogenes* isolates (2019–2023), 272 regional invasive isolates (2009–2019) and 351 carriage isolates (2009–2023). Overall, 72 *emm* types and 147 *emm* subtypes were detected (Supplementary Table S1). In rank order, *emm* types 1, 12, 4, 22 and 89 accounted for 75% among invasive strains and showed partial overlap with the top 5 *emm* types among asymptomatic carriers (*emm* type 12, 1, 6, 75, 4), although their rank differed by time period (Figure 2A). The proportion of *emm*1 was significantly higher among invasive isolates compared to carriage isolates overall (40% vs 15%) but also for each time period, reflecting the known virulent nature of this *emm* type (Figure 2, Supplementary Figure S1). Subgroup analysis of *emm*1 proportions among children below 5 years of age showed a similarly skewed distribution (43% in invasive vs 14% in carriage). In addition to *emm*1, *emm*3 and *emm*22 had higher odds ratios for invasive infection than asymptomatic carriage (Supplementary Table S2).

**Figure 2.**
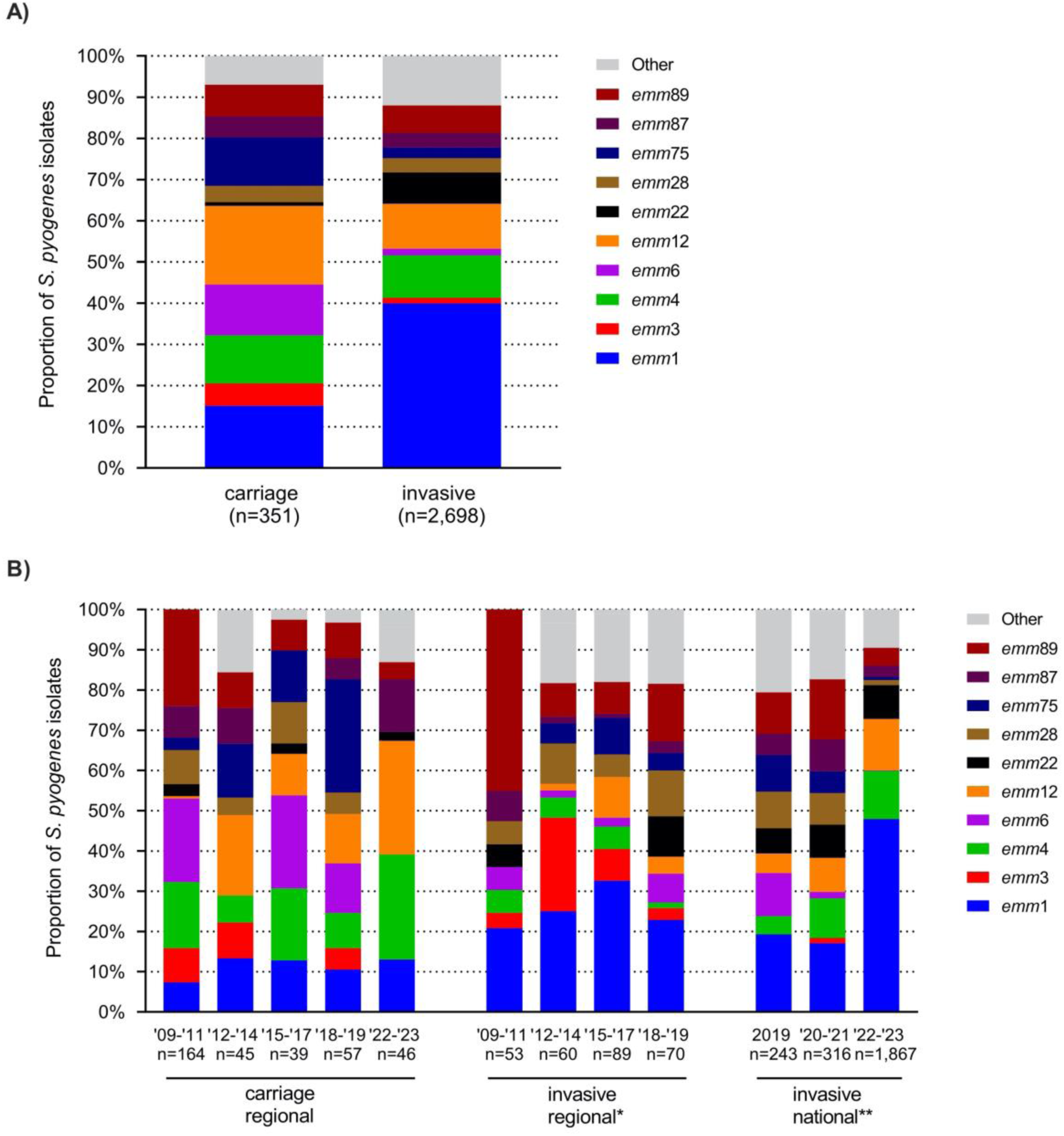
Relative proportion of predominant *emm* types among carriage and invasive *S. pyogenes* isolates in the Netherlands, 2009-2023. Relative proportion of predominant *emm* types among asymptomatic *S. pyogenes* carriers and patients with invasive group A streptococcal infection (A), subdivided by time period (B). This includes 351 carriage isolates from naso- and oropharyngeal swabs from healthy individuals (2009-2023), 272 regional invasive isolates from blood or cerebrospinal fluid (2009-2019), and 2,426 national invasive isolates from any sterile body compartment or non-sterile site with clinical iGAS (2019-2023). *Emm* types with a prevalence of ≥ 10% are included in the figures.

Temporal analysis of *emm* distributions revealed a correlation between the national upsurge of notifiable iGAS manifestations in 2022-2023 and a marked relative and absolute increase of *emm*1.0 among national iGAS strains, from 18 (18%) of 100 isolates in January-March 2022 to 388 (58%) of 670 isolates in January-March 2023 (Figure 3A, B; Fisher’s exact, p<0.0001). The proportion of *emm*1, including subtype *emm*1.0, in iGAS isolates was relatively stable in preceding years (2009-2021; Figure 2B). Strikingly, the post-COVID-19 expansion of *emm*1.0 among iGAS patients was not reflected by higher *emm*1.0 prevalence among carriage isolates collected in the same period: 14% (6/44 isolates) in December 2022 through February 2023, compared to 15% (47/304 isolates) in 2009-2019 (Figure 2B). In addition to *emm*1.0, we observed an emergence and subsequent expansion of a novel *emm*1 subtype, *emm*1.134, among invasive strains in (post-)COVID-19 pandemic years. This new subtype, characterized by four lineage-specific SNPs (Supplementary Table S3), was first detected among invasive isolates in 2020. The *emm* gene of *emm*1.134 differed from *emm*1.0 by a single base pair at position 1683453 of MGAS5005 on the reverse strand, resulting in an amino acid change of asparagine to lysine at position 46 (reading from 5’ to 3’ on the reverse strand). *Emm*1.134 was not detected in 2021, but reappeared in 2022, comprising 29 of 1169 (2.5%) isolates and already 41 (5.6%) of 727 isolates received between Q1 2023 (Figure 3C). Among all iGAS isolates received in 2023, *emm*1.134 ranked 4^th^ as the most prevalent *emm* subtype, after *emm*1.0, *emm* 12.0 and *emm* 4.0. *Emm*1.134 was not detected in carriage isolates (Figure 3D).

**Figure 3.**
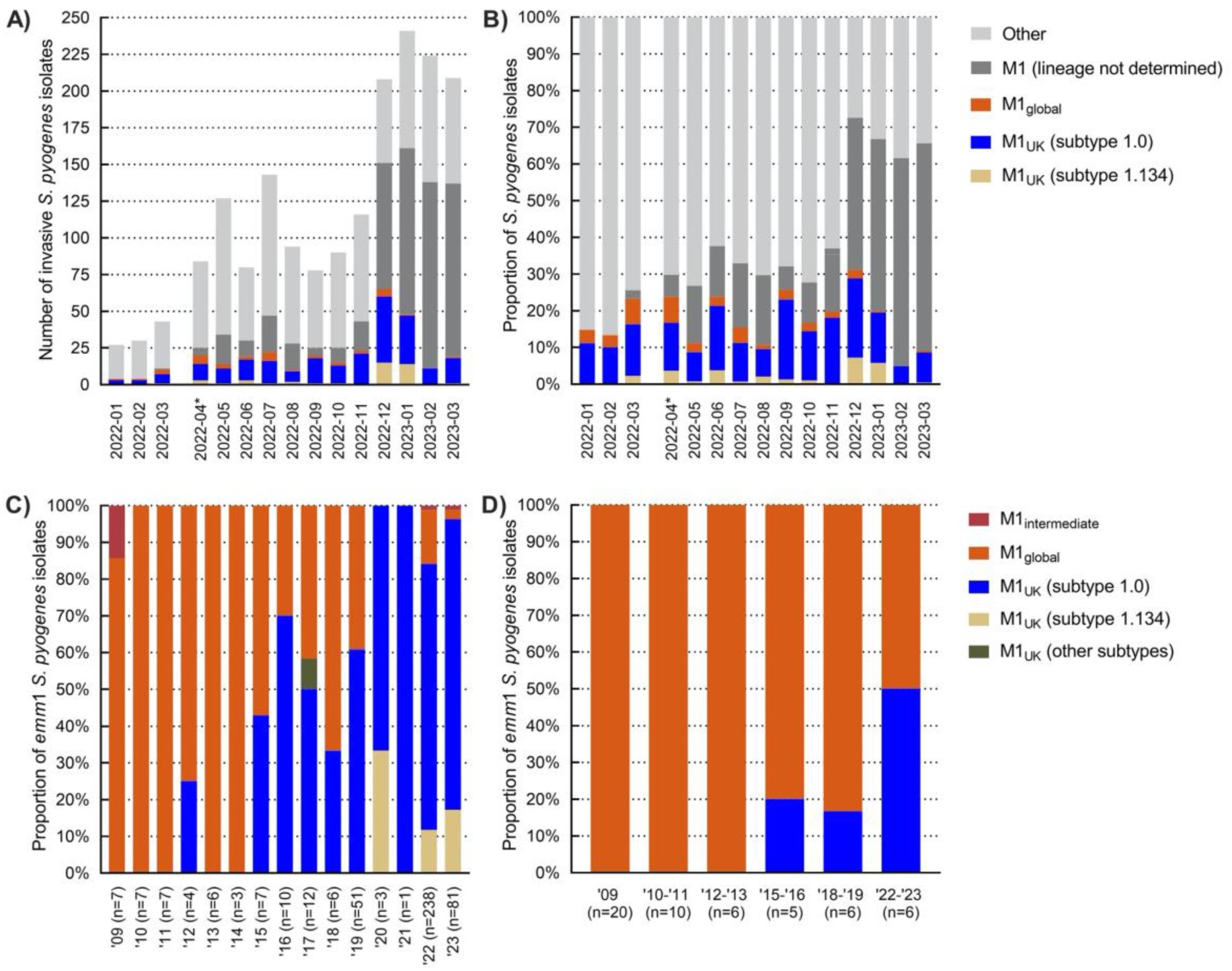
Prevalence of *emm1* subtypes in the Netherlands, 2009-2023. Number (A) and proportion (B) of *emm*1 subtypes among 1,794 invasive strains in 2022-2023 and 444 invasive (C) and 53 carriage (D) *emm*1 *S. pyogenes* strains in 2009-2023. *From May 2022, the number of typed isolates has strongly increased since isolates were received from all medical microbiology laboratories following a national request by the NRLBM and RIVM.

To investigate whether the *emm*1 expansion among iGAS patients was related to a further expansion or diversification of M1_UK_, we sequenced 501 *emm*1 isolates: 367 of 995 (37%) national invasive *emm*1 isolates (2019-2023), all 81 regional *emm*1 invasive isolates (2009-2019) and all 53 *emm*1 carriage isolates (2009-2023). Sequences from two national and two regional invasive isolates were excluded due to low assembly quality. Mapping sequence reads of the remaining 497 isolates against the M1_global_ reference genome (MGAS5005) revealed that the earliest Dutch M1_UK_ isolate, based on presence of 27 lineage-defining SNPs, was detected in a blood culture from 2012. The M1_UK_ lineage became dominant (>50%) among invasive *emm*1 isolates in 2016 (Figure 3C). In the post-COVID-19 years (2022-2023), the proportion of M1_UK_ versus M1_global_ among invasive isolates increased further from 72% (13/18) in Q1 2022 to 96% in Q1 2023 (74/77) (Figure 3C). In contrast, M1_global_ was still highly prevalent (83%, 5 of 6 isolates) among asymptomatic carriers in 2018-2019 and comprised 50% (3 of 6) of *emm*1 carriage isolates during next sampling in winter 2022-2023 (Figure 3D). Notably, all 43 sequenced *emm*1.134 invasive strains belonged to the M1_UK_ lineage based on the conserved presence of the 27 SNPs (1/3 in 2020, 0/1 in 2021, 28/137 in 2022 and 14/82 of sequenced *emm*1 isolates in 2023). Other M1_UK_ isolates were subtype *emm*1.0, except for a single *emm*1.3 in 2017 and one *emm*1.127 in 2019. Five intermediate isolates (based on the presence of 22-25 of the M1_UK_ lineage specific SNPs (11, 17) were detected in 2009, 2022 and 2023.

Phylogenetic comparison showed that M1_UK_ separated into two approximate groups: a clade of highly clonal 2022-2023 isolates and a clade consisting of three smaller distinct branches, including the *emm*1.134 subtype strains (Figure 4; Supplementary Figure S2). Clade-specific SNPs are listed in Supplementary Table S3. Clade separation was not related to the source of *S. pyogenes* isolation (blood, CSF or other sterile body compartments). Although the M1_DK_ lineage was not detected during the 2022/2023 iGAS surge, 15 M1_global_ isolates from 2018-2019 were identified as suspected M1_DK_ based on the presence of 13-14 of the previously reported 15 M1_DK_ lineage specific SNPs (Figure 4). The novel emergent M1_UK_ clades detected in 2022-2023 in the UK (24) could only be identified in two of our sequenced *emm*1 isolates based on presence of clade-specific SNPs. Analysis of the accessory genome showed that *spd1* and *speC* were acquired by 9% (46/497) of all isolates, and 100% (15/15) of suspected M1_DK_ isolates. Other DNAse- and superantigen-encoding genes including *smeZ*, *spd3*, *speA*, *speB*, *speG* and *speJ* were present in 100% of isolates, whereas superantigen gene *ssa* was only detected in a single M1_UK_ isolate. Regulatory genes were screened, and nonsynonymous mutations were found in *covR* in 2.3% (3/132) of M1_global_ and 2.7% (8/296) of M1_UK_ isolates (Fisher’s exact, p>0.05), and in *covS* in 9% (12/132) of M1_global_ and 6.4% (19/296) of M1_UK_ isolates (Fisher’s exact, p>0.05) (Supplementary Table S4). Mutations in these genes did not cluster with a single clade (Figure 4). Proportions of nonsynonymous mutations in other regulatory genes did also not differ between M1_global_ or M1_UK_ strains nor did they show clustering to specific clades (data not shown).

**Figure 4.**
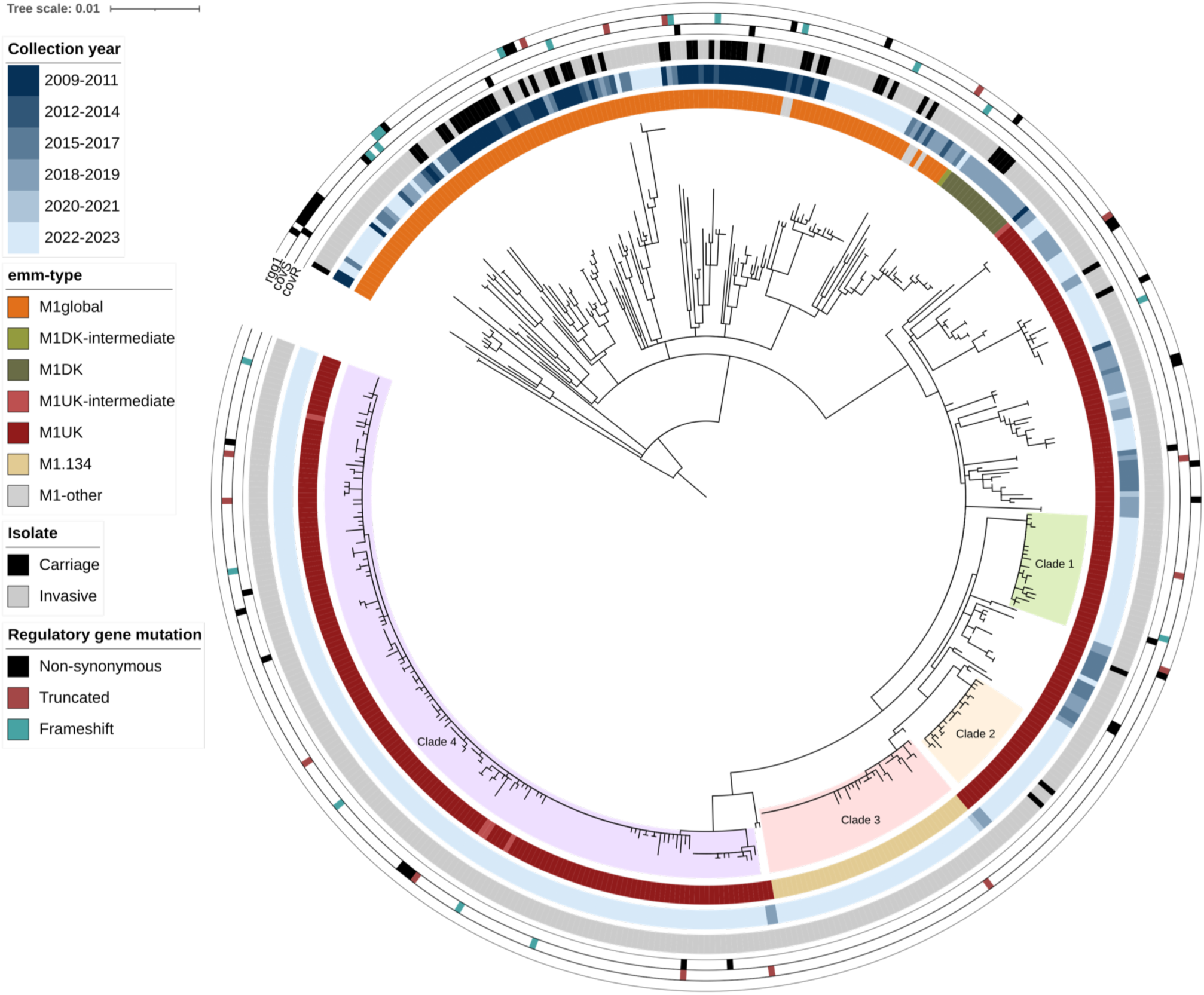
Phylogenetic analysis of invasive and carriage *emm*1 isolates, 2009-2023. Maximum-likelihood phylogenetic tree from the core genome SNP alignment of 444 invasive and 53 carriage Dutch *emm*1 isolates from 2009-2023 to MGAS5005 reference genome (NC_007297.2). The tree was rooted on MGAS5005 and colored rings outside the tree represent year of isolation, *emm* type, isolate and mutations in regulatory genes *covR*, *covS* and *rgg1*. The branches of four distinct clades of recent (2022-2023) M1_UK_ isolates are highlighted within the tree.

## Discussion

The exceptionally high iGAS incidence between March 2022 and March 2023 in the Netherlands coincided with marked expansion of *emm*1 among iGAS isolates. Tracking *emm*1 evolution among invasive isolates indicated that the M1_UK_ lineage was first detected in our country in 2012, became dominant in 2016, and replaced M1_global_ during the steep iGAS increase in late 2022-2023. Four new successful M1_UK_ clades with remarkable clonal stability were detected during the surge, indicating rapid spread. The *emm*1 increase was not reflected in asymptomatic carriers (nor in carriage rates, *emm*1 proportions or M1_UK_ expansion), supporting the hypothesis that a more virulent lineage contributed to the outbreak in our country.

Emergence of lineages with increased pathogenicity and fitness may result in a higher secondary attack rate. Such changes have important implications for the rationale of preventive public health strategies such as antibiotic prophylaxis for close contacts of iGAS patients, since it would lower the number-needed-to-treat. Importantly, UK researchers showed that the expansion of M1_UK_ was detected both in invasive isolates and non-invasive isolates from patients with pharyngitis (24), with increased incidences of scarlet fever prior to the iGAS surge (11). This points to an important role of non-invasive GAS infections, including scarlet fever, in the occurrence of iGAS. Indeed, this is supported by previous studies showing a higher iGAS risk among household contacts of scarlet fever cases (25), and an increased risk of puerperal fever after contact with patients with possible non-invasive GAS (26).

Increases in scarlet fever have not been noted in other European countries, but may be missed since scarlet fever surveillance is not present in all countries. It highlights the importance of surveillance systems that include all clinical GAS manifestations to monitor epidemic clones and guide iGAS control by identification of target groups for prophylactic measures.

An important virulence characteristic of M1_UK_ is increased SpeA production compared to its ancestor M1_global_. SpeA is usually the only phage-encoded superantigen in contemporary *emm*1 *S. pyogenes*, but recent *emm*1 isolates from Denmark (M1_DK_) and Australia (M1_UK_) also carried *speC* and *spd1* (12, 15). The M1_DK_ lineage was present in the Netherlands but not as a driver of the surge in our country. Also the novel emergent M1_UK_ clades detected in 2022- 2023 in the UK (24) were only identified in two of our isolates. It seems that the post-COVID- 19 surge of *emm*1 occurred in a country-specific fashion with the evolution of unique clades in the case of Denmark, the UK, and the Netherlands. Remarkably, the steep iGAS increase occurred in the Netherlands in 2022/2023 whereas the M1_UK_ lineage was already dominant among iGAS since 2016. Acquisition of *speC* and *spd1* was only detected in a small proportion of our *emm*1 isolates, and no SNP profile was shared amongst all Dutch post-COVID-19 *emm*1 iGAS isolates that could fully explain the sudden shift in pathogenicity. Genomic rearrangement resulting in increased transcription of certain virulence factors could also influence lineage-specific pathogenicity and fitness. It would be of interest to explore whether such events could be underlying the clonal expansion of the new successful M1_UK_ clades in our country.

Several countries reported increased iGAS frequencies after COVID-19 lockdowns but (3–5, 12, 27–29), though not all countries reported emergence of specific *emm* types or variants, e.g. Switzerland (30). During 2020-2021, community transmission of all pathogens was particularly low and recovered after lifting COVID-19 social restrictions in 2022. It is posited that during this phase, *emm* type-specific transmission characteristics may have provided a competitive advantage for enhanced spread, such as a stronger reliance on respiratory transmission routes rather than transmission by direct contact between individuals (24). Furthermore, reduced GAS exposure between 2020-2021 may have lowered population immunity against *S. pyogenes*, creating a larger pool of susceptible individuals, particularly among young children. Based on lower circulation of non-*emm*1 *speA-*carrying *emm* types (*emm*3, *emm*6) in the years before COVID-19 lockdowns, we postulate that also *speA*-specific immunity may have been low in post-COVID-19 years. Finally, the post-COVID-19 resurgence of iGAS-associated viruses partly contributed to the iGAS upsurge (10), especially in progression to severe lower respiratory tract infection with pleural empyema (24). Together, we conclude that the unique circumstances of a potentially more susceptible population by lower pathogen-specific immunity and high incidences of predisposing viral infections, in combination with the virulent and fit M1_UK_ lineage already dominant in our country in pre-pandemic years, has coalesced in optimal conditions for extensive expansion of M1_UK_ after COVID-19 restrictions were lifted.

Our study has several limitations. No isolates from non-invasive isolates were included in this study, and therefore we are unfortunately not informed on changes in the epidemiology of non-invasive *S. pyogenes* infections in our country. The sample of carriage isolates was limited to young children and their parents, possibly resulting in skewed *emm* type distributions, since household members are more likely to be colonized by the same *emm* type and because the sampled age groups did not provide a full representation of ages of patients with invasive disease. Carriage isolates were limited to regional sampling, whereas the majority of invasive strains (from 2019 onwards) were collected nationally. Despite these limitations, this is the first study that includes a large set of *S. pyogenes* isolates from asymptomatic carriers collected during the same extended time period (2009–2023) as invasive disease isolates, allowing us to investigate whether changes in carriage rates or carried *emm* types contributed to the iGAS upsurge.

In conclusion, the 2022-2023 iGAS surge in the Netherlands is linked to clonal replacement and suspected increased virulence of four new successful M1_UK_ clades, possibly augmented by lower *S. pyogenes* population immunity and increased circulation of iGAS- predisposing viruses after lifting COVID-19 restrictions. Asymptomatic carriers do not seem to represent a major reservoir of invasive infections attributed to this lineage. The rise of more virulent clades may have implications for secondary attack rates and the rationale for post-exposure antibiotic prophylaxis for close contacts of index patients, supporting the importance of vigilant monitoring of *S. pyogenes* infections and timely detection of epidemic clones. Future studies that include the complete clinical spectrum of *S. pyogenes* infections are warranted to identify transmission routes and possible public health interventions to reduce iGAS burden.

## Supporting information

Supplementary data isolates

## Data Availability

Whole genome sequence data will be publically available through the PubMLST database: https://pubmlst.org/organisms/streptococcus-pyogenes

## Acknowledgements.

We thank all medical microbiological laboratories for submitting iGAS isolates and the NRLBM technicians for typing all isolates.

## Author contributions

LWR, SMTV, BdG, BJMV, AvdE and NMvS contributed to funding acquisition, data collection and interpretation. WCMBK, MAvH, NYR, AWM contributed to data collection and interpretation. LWR, MD, BCLvdP, BdG analyzed the data and produced figures and tables. LWR, MD and NMvS wrote the manuscript. NMvS supervised the study. All authors provided

