## Supplemental information for "Nationwide upsurge in invasive disease in the context of longitudinal surveillance of carriage and invasive *Streptococcus pyogenes* 2009-2023, the Netherlands: a molecular epidemiological study"

**Supplement**

**Supplementary methods**

*Emm typing and whole genome sequencing*

*Emm*-genotyped was performed by conventional PCR amplification and subsequent Sanger sequencing of the 180-bp hypervariable domain of the *emm* gene according to the CDC protocol (1). Next-generation sequencing was performed using Illumina short-read technology (Microbes NG, Birmingham, UK and in-house core facility Amsterdam UMC).

*Genome assembly*

Raw reads were processed with Trimmomatic v0.39 (2) to remove adaptor sequences and bases of insufficient quality. Trimmed reads were *de novo* assembled into scaffolds with SPAdes v3.15.5 and assembly quality was assessed with QUAST v5.0.2 (3).

*Core SNP phylogenetic analysis*

A core SNP alignment was produced by calling SNPs against reference genome MGAS5005 (NCBI accession: NC_007297) with snippy v4.6.0 (<https://github.com/tseemann/snippy>) using default parameters. Sites of recombination were detected and masked with Gubbins v2.3.4 (4) and a phylogenetic inference by maximum likelihood was performed on the subsequent alignment by IQ-TREE (5) using the ‘GTR +G +I’ substitution model. Phylogenetic tree was visualized using iTOL v6.9 (6).

Snippy was also utilised to classify clade-specific SNPs into synonymous, non-synonymous or frameshift mutations. A SNP distance matrix was determined with snp-dists v0.8.2 (https://github.com/tseemann/snp-dists) and visualised with R (7) package pheatmap .

*Genes of interest analysis*

The presence of DNAse, superantigen and regulatory genes was determined with BLASTN v2.15.0 (8), gene sequences were extracted using faidx (9) and translated into amino acid sequences with MACSE program ‘translateNT2AA’ (10). Amino acid sequences were compared against MGAS5005 to determine non-synonymous mutations. Resistance genes were screened for with ABRicate v1.0.1 (<https://github.com/tseemann/abricate>).

**Supplementary Figure S1. Relative proportion of predominant *emm* types among 3,049 *S. pyogenes* isolates collected between 2009-2023, the Netherlands.**

Relative proportion of predominant *emm* types among 351 regional carriage (Noord-Holland region, 2009-2023), 272 regional invasive (Utrecht region, 2009-2019) and 2,426 national invasive (2019-2023) *S. pyogenes* isolates. Each point indicates the proportion of carriage isolates (on the x-axis) and invasive isolates (on the y-axis) by *emm* type. The dotted line indicates where the two proportions are equal. Red color indicates that *emm* types were significantly more common among invasive isolates by binary logistic regression adjusted for time period and age group. *Emm* types indicated with a black dot did not reach statistical significance.

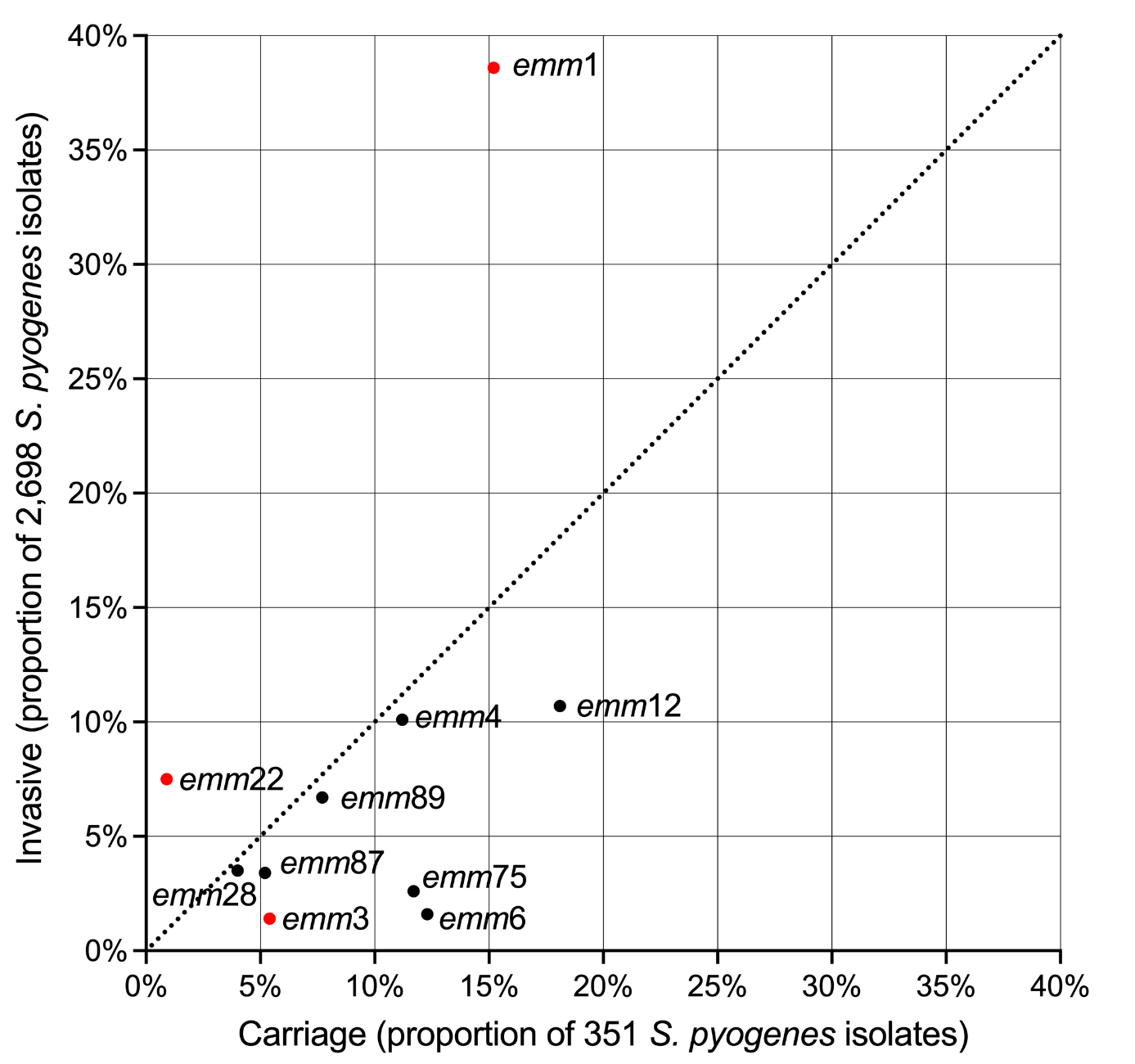

**Supplementary Figure S2. SNP distance analysis of *emm1* genomes from the Netherlands between 2009-2023.**

SNP distance matrix based on 2028 core SNPs between 497 *emm1* genomes. Colors in the heatmap indicate the number of SNPs between isolates. with highly similar isolates in blue and shifting towards red as number of SNPs increases. *Emm* type and M1_UK_-specific clades are visualized by the top dendrogram. which has been clustered with hierarchical clustering from R package pheatmap.

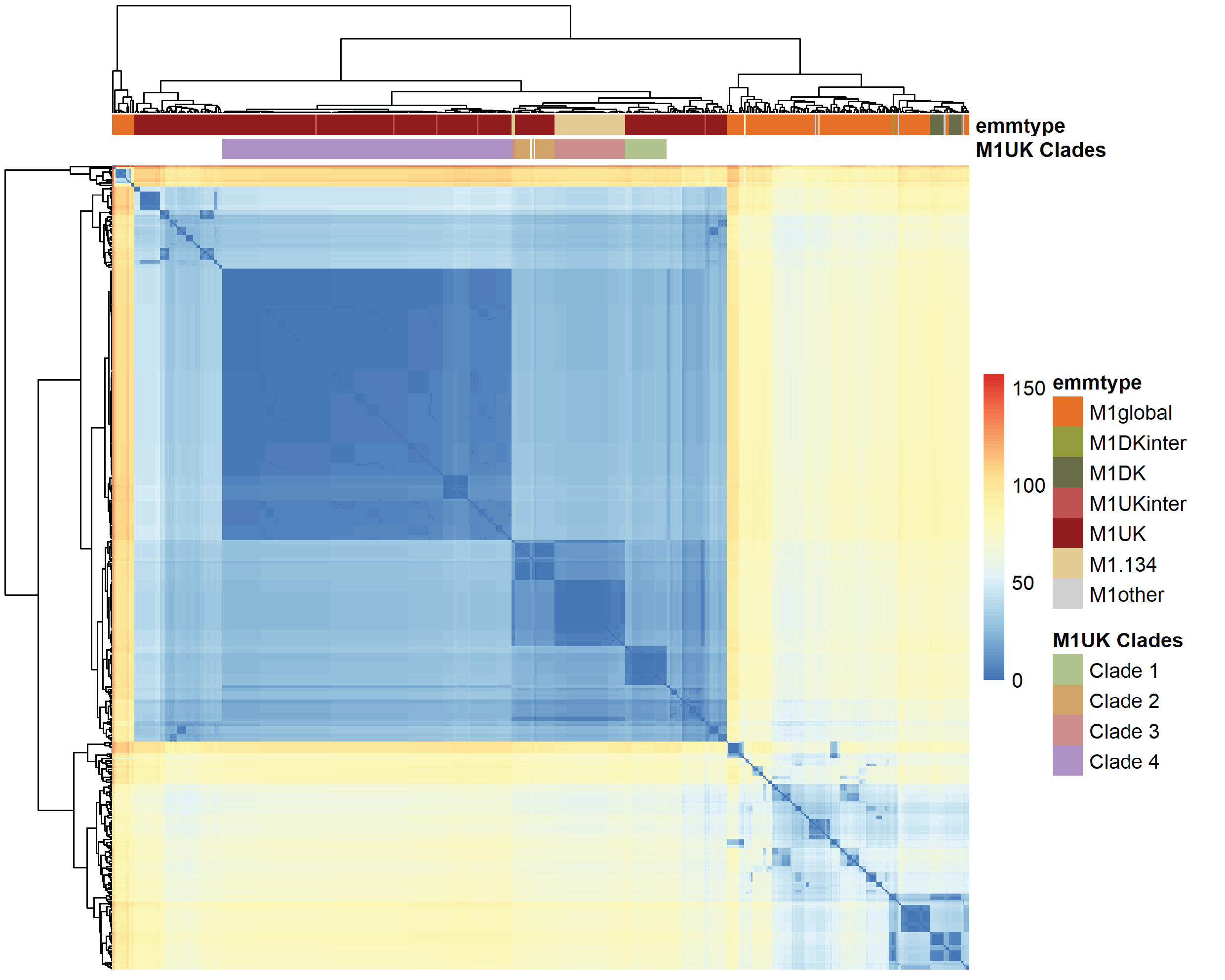

**Supplementary Table S1. *Emm* (sub)types of national invasive strains (n=2,426), regional invasive *S. pyogenes* strains (n=272) and carriage strains (n=351) in the Netherlands, 2009-2023**

| Subtype | Cluster | Carriage regional  (2009-2023) | | Invasive regional (2009-2019) | | Invasive national  (2019-2023) | | Total |  |
| --- | --- | --- | --- | --- | --- | --- | --- | --- | --- |
| 1.0 | A-C3 | 52 | 14.8% | 78 | 28.7% | 899 | 37.1% | 1029 | 33.7% |
| 1.100 | A-C3 | 0 |  | 0 |  | 1 | 0.0% | 1 | 0.0% |
| 1.109 | A-C3 | 0 |  | 0 |  | 1 | 0.0% | 1 | 0.0% |
| 1.120 | A-C3 | 0 |  | 0 |  | 1 | 0.0% | 1 | 0.0% |
| 1.127 | A-C3 | 0 |  | 0 |  | 2 | 0.1% | 2 | 0.1% |
| 1.134 | A-C3 | 0 |  | 0 |  | 71 | 2.9% | 71 | 2.3% |
| 1.15 | A-C3 | 0 |  | 0 |  | 2 | 0.1% | 2 | 0.1% |
| 1.152 | A-C3 | 0 |  | 0 |  | 1 | 0.0% | 1 | 0.0% |
| 1.18 | A-C3 | 0 |  | 0 |  | 1 | 0.0% | 1 | 0.0% |
| 1.19 | A-C3 | 0 |  | 0 |  | 1 | 0.0% | 1 | 0.0% |
| 1.25 | A-C3 | 1 | 0.3% | 0 |  | 3 | 0.1% | 4 | 0.1% |
| 1.29 | A-C3 | 0 |  | 0 |  | 2 | 0.1% | 2 | 0.1% |
| 1.3 | A-C3 | 0 |  | 1 | 0.4% | 7 | 0.3% | 8 | 0.3% |
| 1.33 | A-C3 | 0 |  | 0 |  | 1 | 0.0% | 1 | 0.0% |
| 1.4 | A-C3 | 0 |  | 2 | 0.7% | 0 |  | 2 | 0.1% |
| 1.6 | A-C3 | 0 |  | 0 |  | 1 | 0.0% | 1 | 0.0% |
| 1.87 | A-C3 | 0 |  | 0 |  | 1 | 0.0% | 1 | 0.0% |
| 2.0 | E4 | 6 | 1.7% | 1 | 0.4% | 4 | 0.2% | 11 | 0.4% |
| 3.1 | A-C5 | 16 | 4.6% | 27 | 9.9% | 0 |  | 43 | 1.4% |
| 3.155 | A-C5 | 1 | 0.3% | 0 |  | 0 |  | 1 | 0.0% |
| 3.17 | A-C5 | 0 |  | 1 | 0.4% | 0 |  | 1 | 0.0% |
| 3.23 | A-C5 | 0 |  | 1 | 0.4% | 0 |  | 1 | 0.0% |
| 3.61 | A-C5 | 0 |  | 1 | 0.4% | 0 |  | 1 | 0.0% |
| 3.93 | A-C5 | 2 | 0.6% | 4 | 1.5% | 4 | 0.2% | 10 | 0.3% |
| 4.0 | E1 | 41 | 11.7% | 11 | 4.0% | 253 | 10.5% | 305 | 10.0% |
| 4.14 | E1 | 0 |  | 0 |  | 1 | 0.0% | 1 | 0.0% |
| 4.19 | E1 | 0 |  | 0 |  | 8 | 0.3% | 8 | 0.3% |
| 4.26 | E1 | 0 |  | 0 |  | 2 | 0.1% | 2 | 0.1% |
| 4.28 | E1 | 0 |  | 0 |  | 1 | 0.0% | 1 | 0.0% |
| 4.9 | E1 | 0 |  | 0 |  | 1 | 0.0% | 1 | 0.0% |
| 5.199 | M5 | 0 |  | 1 | 0.4% | 0 |  | 1 | 0.0% |
| 5.23 | M5 | 1 | 0.3% | 1 | 0.4% | 1 | 0.0% | 3 | 0.1% |
| 5.44 | M5 | 0 |  | 0 |  | 1 | 0.0% | 1 | 0.0% |
| 5.46 | M6 | 1 | 0.3% | 0 |  | 0 |  | 1 | 0.0% |
| 5.75 | M5 | 1 | 0.3% | 0 |  | 0 |  | 1 | 0.0% |
| 6.0 | M6 | 20 | 5.7% | 8 | 2.9% | 24 | 1.0% | 52 | 1.7% |
| 6.19 | M6 | 1 | 0.3% | 0 |  | 0 |  | 1 | 0.0% |
| 6.4 | M6 | 20 | 5.7% | 3 | 1.1% | 7 | 0.3% | 30 | 1.0% |
| 6.5 | M6 | 2 | 0.6% | 0 |  | 0 |  | 2 | 0.1% |
| 6.98 | M6 | 0 |  | 0 |  | 1 | 0.0% | 1 | 0.0% |
| 8.0 | E4 | 0 |  | 0 |  | 5 | 0.2% | 5 | 0.1% |
| 9.0 | E3 | 4 | 1.1% | 5 | 1.8% | 5 | 0.2% | 14 | 0.5% |
| 9.4 | E3 | 0 |  | 1 | 0.4% | 1 | 0.0% | 2 | 0.1% |
| 11.0 | E6 | 1 | 0.3% | 1 | 0.4% | 28 | 1.2% | 30 | 1.0% |
| 11.31 | E6 | 0 |  | 0 |  | 1 | 0.0% | 1 | 0.0% |
| 12.0 | A-C4 | 56 | 16.0% | 13 | 4.8% | 215 | 8.9% | 284 | 9.3% |
| 12.19 | A-C4 | 0 |  | 0 |  | 2 | 0.1% | 2 | 0.1% |
| 12.32 | A-C4 | 0 |  | 0 |  | 1 | 0.0% | 1 | 0.0% |
| 12.37 | A-C4 | 11 | 3.1% | 3 | 1.1% | 58 | 2.4% | 72 | 2.4% |
| 12.55 | A-C4 | 0 |  | 0 |  | 1 | 0.0% | 1 | 0.0% |
| 12.68 | A-C4 | 0 |  | 0 |  | 1 | 0.0% | 1 | 0.0% |
| 12.7 | A-C4 | 2 | 0.6% | 0 |  | 0 |  | 2 | 0.1% |
| 14.15 | Y | 0 |  | 0 |  | 1 | 0.0% | 1 | 0.0% |
| 14.3 | Y | 0 |  | 0 |  | 1 | 0.0% | 1 | 0.0% |
| 18.21 | Y | 0 |  | 1 | 0.4% | 0 |  | 1 | 0.0% |
| 18.23 | Y | 0 |  | 0 |  | 1 | 0.0% | 1 | 0.0% |
| 22.0 | E4 | 2 | 0.6% | 7 | 2.6% | 194 | 8.0% | 203 | 6.7% |
| 22.1 | E4 | 0 |  | 0 |  | 1 | 0.0% | 1 | 0.0% |
| 22.21 | E4 | 0 |  | 0 |  | 2 | 0.1% | 2 | 0.1% |
| 22.3 | E4 | 1 | 0.3% | 0 |  | 0 |  | 1 | 0.0% |
| 24.9 | Y | 0 |  | 0 |  | 1 | 0.0% | 1 | 0.0% |
| 25.1 | E3 | 0 |  | 0 |  | 2 | 0.1% | 2 | 0.1% |
| 27.0 | E2 | 0 |  | 0 |  | 2 | 0.1% | 2 | 0.1% |
| 27.6 | E2 | 0 |  | 0 |  | 4 | 0.2% | 4 | 0.1% |
| 28.0 | E4 | 14 | 4.0% | 20 | 7.4% | 70 | 2.9% | 104 | 3.4% |
| 28.14 | E4 | 0 |  | 1 | 0.4% | 1 | 0.0% | 2 | 0.1% |
| 28.5 | E4 | 0 |  | 1 | 0.4% | 1 | 0.0% | 2 | 0.1% |
| 29.2 | M29 | 0 |  | 0 |  | 1 | 0.0% | 1 | 0.0% |
| 33.0 | D4 | 0 |  | 1 | 0.4% | 4 | 0.2% | 5 | 0.2% |
| 43.4 | D4 | 0 |  | 0 |  | 1 | 0.0% | 1 | 0.0% |
| 44.0 | E3 | 0 |  | 5 | 1.8% | 12 | 0.5% | 17 | 0.6% |
| 48.1 | E6 | 0 |  | 1 | 0.4% | 4 | 0.2% | 5 | 0.2% |
| 49.0 | E3 | 0 |  | 0 |  | 2 | 0.1% | 2 | 0.1% |
| 49.3 | E3 | 0 |  | 3 | 1.1% | 5 | 0.2% | 8 | 0.3% |
| 49.8 | E3 | 0 |  | 0 |  | 8 | 0.3% | 8 | 0.3% |
| 52.1 | D4 | 0 |  | 0 |  | 1 | 0.0% | 1 | 0.0% |
| 53.1 | D4 | 0 |  | 0 |  | 2 | 0.1% | 2 | 0.1% |
| 53.12 | D4 | 0 |  | 0 |  | 2 | 0.1% | 2 | 0.1% |
| 56.2 | D4 | 0 |  | 0 |  | 1 | 0.0% | 1 | 0.0% |
| 56.4 | D4 | 0 |  | 0 |  | 1 | 0.0% | 1 | 0.0% |
| 57.4 | Y | 1 | 0.3% | 0 |  | 0 |  | 1 | 0.0% |
| 58.0 | E3 | 0 |  | 0 |  | 8 | 0.3% | 8 | 0.3% |
| 58.22 | E3 | 0 |  | 0 |  | 1 | 0.0% | 1 | 0.0% |
| 60.11 | E1 | 0 |  | 0 |  | 1 | 0.0% | 1 | 0.0% |
| 65.4 | E6 | 0 |  | 0 |  | 1 | 0.0% | 1 | 0.0% |
| 66.0 | E2 | 0 |  | 0 |  | 1 | 0.0% | 1 | 0.0% |
| 66.1 | E2 | 0 |  | 0 |  | 2 | 0.1% | 2 | 0.1% |
| 67.0 | E6 | 0 |  | 0 |  | 1 | 0.0% | 1 | 0.0% |
| 68.0 | E2 | 0 |  | 0 |  | 1 | 0.0% | 1 | 0.0% |
| 68.3 | E2 | 0 |  | 1 | 0.4% | 2 | 0.1% | 3 | 0.1% |
| 73.0 | E4 | 0 |  | 0 |  | 5 | 0.2% | 5 | 0.2% |
| 74.0 | Y | 0 |  | 0 |  | 1 | 0.0% | 1 | 0.0% |
| 75.0 | E6 | 41 | 11.7% | 17 | 6.3% | 52 | 2.1% | 110 | 3.6% |
| 76.0 | E2 | 0 |  | 0 |  | 9 | 0.4% | 9 | 0.3% |
| 76.13 | E2 | 0 |  | 0 |  | 2 | 0.1% | 2 | 0.1% |
| 76.17 | E2 | 0 |  | 0 |  | 4 | 0.2% | 4 | 0.1% |
| 76.21 | E2 | 0 |  | 0 |  | 1 | 0.0% | 1 | 0.0% |
| 77.0 | E4 | 1 | 0.3% | 7 | 2.6% | 42 | 1.7% | 50 | 1.6% |
| 78.3 | E1 | 0 |  | 1 | 0.4% | 0 |  | 1 | 0.0% |
| 80.7 | D4 | 0 |  | 0 |  | 1 | 0.0% | 1 | 0.0% |
| 81.0 | E6 | 1 | 0.3% | 1 | 0.4% | 12 | 0.5% | 14 | 0.5% |
| 81.1 | E6 | 0 |  | 1 | 0.4% | 0 |  | 1 | 0.0% |
| 81.2 | E6 | 0 |  | 0 |  | 1 | 0.0% | 1 | 0.0% |
| 81.5 | E6 | 0 |  | 0 |  | 1 | 0.0% | 1 | 0.0% |
| 82.0 | E3 | 2 | 0.6% | 0 |  | 15 | 0.6% | 17 | 0.6% |
| 82.1 | E3 | 0 |  | 0 |  | 1 | 0.0% | 1 | 0.0% |
| 83.1 | D4 | 0 |  | 0 |  | 3 | 0.1% | 3 | 0.1% |
| 84.0 | E4 | 0 |  | 1 | 0.4% | 0 |  | 1 | 0.0% |
| 85.0 | E6 | 0 |  | 0 |  | 3 | 0.1% | 3 | 0.1% |
| 85.1 | E6 | 0 |  | 0 |  | 1 | 0.0% | 1 | 0.0% |
| 87.0 | E3 | 17 | 4.8% | 4 | 1.5% | 90 | 3.7% | 111 | 3.6% |
| 87.16 | E3 | 1 | 0.3% | 0 |  | 0 |  | 1 | 0.0% |
| 88.2 | E4 | 0 |  | 0 |  | 1 | 0.0% | 1 | 0.0% |
| 88.5 | E4 | 0 |  | 1 | 0.4% | 0 |  | 1 | 0.0% |
| 89.0 | E4 | 26 | 7.4% | 26 | 9.6% | 155 | 6.4% | 207 | 6.8% |
| 89.24 | E4 | 1 | 0.3% | 0 |  | 0 |  | 1 | 0.0% |
| 89.42 | E4 | 0 |  | 0 |  | 1 | 0.0% | 1 | 0.0% |
| 90.2 | E2 | 0 |  | 0 |  | 3 | 0.1% | 3 | 0.1% |
| 90.5 | E2 | 0 |  | 0 |  | 1 | 0.0% | 1 | 0.0% |
| 92.0 | E2 | 0 |  | 0 |  | 2 | 0.1% | 2 | 0.1% |
| 93.0 | D4 | 0 |  | 0 |  | 1 | 0.0% | 1 | 0.0% |
| 94.0 | E6 | 0 |  | 0 |  | 5 | 0.2% | 5 | 0.2% |
| 94.1 | E6 | 0 |  | 1 | 0.4% | 2 | 0.1% | 3 | 0.1% |
| 95.0 | - | 0 |  | 0 |  | 3 | 0.1% | 3 | 0.1% |
| 99.6 | E6 | 0 |  | 0 |  | 1 | 0.0% | 1 | 0.0% |
| 102.2 | E4 | 0 |  | 0 |  | 3 | 0.1% | 3 | 0.1% |
| 102.3 | E4 | 0 |  | 1 | 0.4% | 6 | 0.2% | 7 | 0.2% |
| 102.9 | E4 | 0 |  | 0 |  | 1 | 0.0% | 1 | 0.0% |
| 103.0 | E3 | 0 |  | 0 |  | 1 | 0.0% | 1 | 0.0% |
| 104.0 | E2 | 0 |  | 0 |  | 7 | 0.3% | 7 | 0.2% |
| 106.0 | E2 | 0 |  | 0 |  | 1 | 0.0% | 1 | 0.0% |
| 108.10 | D4 | 0 |  | 0 |  | 1 | 0.0% | 1 | 0.0% |
| 118.0 | E3 | 0 |  | 0 |  | 6 | 0.2% | 6 | 0.1% |
| 118.2 | E3 | 0 |  | 1 | 0.4% | 0 |  | 1 | 0.0% |
| 122.2 | Y | 0 |  | 0 |  | 1 | 0.0% | 1 | 0.0% |
| 124.0 | E4 | 0 |  | 1 | 0.4% | 0 |  | 1 | 0.0% |
| 145.1 | - | 0 |  | 0 |  | 1 | 0.0% | 1 | 0.0% |
| 145.5 | - | 0 |  | 1 | 0.4% | 0 |  | 1 | 0.0% |
| 168.0 | E2 | 0 |  | 0 |  | 1 | 0.0% | 1 | 0.0% |
| 169.3 | E4 | 0 |  | 0 |  | 1 | 0.0% | 1 | 0.0% |
| 169.8 | E4 | 0 |  | 0 |  | 1 | 0.0% | 1 | 0.0% |
| 170.0 | E5 | 0 |  | 0 |  | 1 | 0.0% | 1 | 0.0% |
| 177.0 | E6 | 0 |  | 1 | 0.4% | 0 |  | 1 | 0.0% |
| 183.2 | E3 | 0 |  | 0 |  | 2 | 0.1% | 2 | 0.1% |
| 209.0 | E3 | 0 |  | 0 |  | 1 | 0.0% | 1 | 0.0% |
| 236.0 | X | 1 | 0.3% | 0 |  | 0 |  | 1 | 0.0% |
| 264.0 | - | 0 |  | 0 |  | 1 | 0.0% | 1 | 0.0% |
| Unknown | - | 3 | 0.9% | 3 | 1.1% | 6 | 0.2% | 12 | 0.4% |
| Total |  | **351** | **100%** | **272** | **100.0%** | **2426** | **100%** | **3049** | **100%** |

**Supplementary Table S2. *Emm* types associated with invasive infection versus asymptomatic carriage: binary logistic regression analysis.**

Number and proportion of *emm* types (1, 3, 6, 12, 22, 28, 75, 87, 89, other *emm* types), and the odds ratios (OR) with 95% confidence intervals (95% CI) for invasive group A streptococcal infection compared to asymptomatic *S. pyogenes* carriage, calculated by binary logistic regression adjusted for time period of sample collection (2009-2011, 2012-2014, 2015-2017, 2018-2021, 2022-2023) and age group (child, adult). *Emm*4 served as a reference category.

| *Emm* type | Carriage (%) (n=351) | Invasive (%) (n=2,698) | Adjusted OR (95% CI) | p-value |
| --- | --- | --- | --- | --- |
| *Emm*1 | 53 (15.2) | 1076 (40.0) | 4.0 (2.2 - 7.4) | **0** |
| *Emm*3 | 19 (5.5) | 38 (1.4) | 2.5 (1.1 - 6.1) | **0.04** |
| *Emm*6 | 43 (12.4) | 43 (1.6) | 0.5 (0.2 - 1.2) | 0.12 |
| *Emm*12 | 67 (19.3) | 294 (10.9) | 0.8 (0.4 - 1.5) | 0.55 |
| *Emm*22 | 3 (0.9) | 204 (7.6) | 5.4 (1.6 - 27.1) | **0.02** |
| *Emm*28 | 14 (4.0) | 94 (3.5) | 2.4 (0.9 - 6.3) | 0.08 |
| *Emm*75 | 41 (11.8) | 69 (2.6) | 0.5 (0.2 - 1.1) | 0.08 |
| *Emm*87 | 18 (5.2) | 94 (3.5) | 0.5 (0.2 - 1.2) | 0.11 |
| *Emm*89 | 27 (7.8) | 182 (6.8) | 1.5 (0.7 - 3.2) | 0.33 |
| Other *emm* types | 22 (6.3) | 318 (11.8) | 2.2 (1.0 - 4.8) | 0.04 |
| *Emm*4 | 41 (11.8) | 277 (10.3) | Reference category |  |

**Supplementary Table S3. Clade-specific single nucleotide polymorphisms in 4 distinct *emm*1 clades detected by whole genome sequencing in the Netherlands, 2022-2023**

| Clade | MGAS5005 Location | Locus tag (M5005_) | Gene | Product | S/NS | Amino acid change | Ref | SNP/ indel |
| --- | --- | --- | --- | --- | --- | --- | --- | --- |
| Clade 1 M1_UK_ (24 isolates) | 220093 | RS01235 |  | sugar ABC transporter permease | S | Tyr96Tyr | C | T |
|  | 484377 | RS02590 | *thiT* | energy-coupled thiamine transporter ThiT | NS | Leu133Phe | G | A |
|  | 652589 | RS03315 |  | glycerophosphoryl diester phosphodiesterase membrane domain-containing protein | NS | Leu62Phe | A | T |
|  | 739015 | RS03745 | *rfbB* | dTDP-glucose 4.6-dehydratase | NS | Tyr153His | T | C |
|  | 760210 | RS03830 | *hylA* | hyaluronidase | S | Asn591Asn | A | G |
|  | 856215 | RS04340 |  | serine hydroxymethyltransferase | NS | His20Tyr | C | T |
|  | 1193375 | RS06110 |  | amino acid ABC transporter ATP-binding protein | NS | Met79Ile | C | T |
|  | 1573502 | RS07990 | *truA* | tRNA pseudouridine(38-40) synthase TruA | S | Asn227Asn | A | G |
|  | 1699039 | Intergenic |  |  | Intergenic |  | G | A |
| Clade 2 M1_UK­_ (21 isolates) | 140190 | RS00870 |  | V-type ATP synthase subunit A | NS | Gly106Val | G | T |
|  | 301646 | RS01610 |  | DEAD/DEAH box helicase | NS | Ala881Ser | G | T |
|  | 328883 | RS01780 |  | iron ABC transporter permease | NS | Ala128Thr | C | T |
|  | 456537 | RS10035 |  | histidyl-tRNA synthetase | Frame shift | Leu74fs | T | TA |
|  | 548355^*^ | RS02870 | *eno* | surface-displayed alpha-enolase | NS | Arg254Cys | C | T |
|  | 878394 | RS04435 |  | PTS sugar transporter subunit IIC | NS | Gly48Ser | G | A |
|  | 1380619^*^ | RS06990 | *infB* | translation initiation factor IF-2 | NS | Met674Ile | C | T |
|  | 1410699 | RS07160 |  | hypothetical protein | Frame shift | Asn32fs | AT | A |
| Clade 3 M1.134 (43 isolates) | 718396 | RS03625 |  | branched-chain amino acid aminotransferase | S | Tyr124Tyr | C | T |
|  | 810007 | RS04075 |  | lantibiotic immunity ABC transporter MutG family permease subunit | NS | Gly160Asp | G | A |
|  | 871566^†^ | RS04400 |  | hemolysin III family protein | Frame shift | Tyr99fs | T | TAA |
|  | 1028811 | RS05245 |  | extracellular solute-binding protein | NS | Ser233Asn | G | A |
|  | 1683453 | RS08475 | *emm* | M protein | NS | Asn46Lys | A | C |
| Clade 4 M1_UK_ (168 isolates) | 102363 | M5005_RS00645 | *comGA* | competence type IV pilus ATPase ComGA | S | Gly95Gly | T | C |
|  | 229549 | RS01285 | *rpe* | ribulose-phosphate 3-epimerase | S | Gly146Gly | T | G |
|  | 396830^*^ | RS02145 |  | hypothetical protein | NS | Thr52Ile | C | T |
|  | 600386 |  |  |  | Intergenic |  | G | A |
|  | 645318^*^ | RS03290 |  | carbamoyl phosphate synthase small subunit | NS | Thr358Met | C | T |
|  | 717680^*^ | RS03620 | *parC* | DNA topoisomerase IV subunit A | S | Leu746Leu | G | A |
|  | 768422 | RS03860 | *glmM* | phosphoglucosamine mutase | S | Thr337Thr | C | A |
|  | 773188 | Intergenic |  |  | Intergenic |  | T | G |
|  | 913835 | Intergenic |  |  | Intergenic |  | C | T |
|  | 1003656^*^ | RS05075 |  | minor capsid protein | Frame shift | Ile120fs | A | AT |
|  | 1355235^*^ | Intergenic |  |  | Intergenic |  | C | T |
|  | 1415991^‡^ | RS07185 |  | phage/plasmid primase. P4 family | S | Val150Val | G | A |
|  | 1423424^‡^ | Intergenic |  |  | Intergenic |  | C | T |
|  | 1649676^§^ | RS08330 | *slc1* | adhesin Scl1 | NS | Gly31Val | C | A |
|  | 1730494 | RS08700 | *ahpF* | alkyl hydroperoxide reductase subunit F | NS | Met398Val | A | G |
|  | 1781667 | RS08915 | *aspS* | aspartate--tRNA ligase | NS | Glu568Gly | T | C |
|  | 1795728^*^ | RS08990 |  | YhgE/Pip domain-containing protein | NS | Asn264Asp | T | C |
|  | 1818613^†^ | Intergenic |  |  | Intergenic |  | T | TG |

Clade-specific SNP annotations against reference strain (Ref) MGAS5005 (NC_007297.2) determined with Snippy. S/NS refers to synonymous and non-synonymous mutations, respectively.

*Also present in the two most related isolates to the clade.

†Missing from one isolate in the clade.

‡Missing from <3% of isolates in the clade.

§Missing from <10% of isolates in the clade.

**Supplementary Table S4. Mutations in regulatory genes *covR*, *covS*, and *rgg* among different *emm*1 subtypes**

|  | Non synonymous  (n, %) | Frameshift  Truncated  Non-functional  (n, %) | # isolates |
| --- | --- | --- | --- |
| M1_global_ | 3 (2.3) | 0 | 132 |
| M1_DKinter_ | 0 | 0 | 1 |
| M1_DK_ | 0 | 0 | 15 |
| M1_UKinter_ | 0 | 0 | 5 |
| M1_UK_ | 8 (2.7) | 0 | 296 |
| M1.134 | 0 | 0 | 43 |
| M1other | 0 | 0 | 5 |

A) ***Emm*1 isolates with mutations in *covR***

|  | Non synonymous (n, %) | Frameshift  Truncated  Non-functional  (n, %) | # isolates |
| --- | --- | --- | --- |
| M1_global_ | 2 (1.5) | 10 (7.8) | 132 |
| M1_DKinter_ | 0 | 0 | 1 |
| M1_DK_ | 0 | 0 | 15 |
| M1_UKinter_ | 0 | 0 | 5 |
| M1_UK_ | 8 (2.7) | 11 (3.7) | 296 |
| M1.134 | 1 (2.3) | 0 | 43 |
| M1other | 0 | 1 (20) | 5 |

B) ***Emm* isolates with mutations in *covS***
C) ***Emm* isolates with mutations in *rgg***

|  | Non synonymous  (n, %) | Frameshift  Truncated  Non-functional  (n, %) | # isolates |
| --- | --- | --- | --- |
| M1_global_ | 11 (8.3) | 2 (1.5) | 132 |
| M1_DKinter_ | 0 | 0 | 1 |
| M1_DK_ | 1 (6.7) | 0 | 15 |
| M1_UKinter_ | 0 | 0 | 5 |
| M1_UK_ | 8 (2.7) | 0 | 296 |
| M1.134 | 0 | 0 | 43 |
| M1other | 0 | 0 | 5 |
